# Children Living with Disabilities Are Absent From Severe Malnutrition Guidelines

**DOI:** 10.1101/2021.09.24.21264078

**Authors:** Magdalena Engl, Paul Binns, Indi Trehan, Natasha Lelijveld, Chloe Angood, Marie McGrath, Nora Groce, Marko Kerac

**Author notes:** Corresponding author: Magdalena Engl, Keppel St, London WC1E 7HT.

## Abstract

**Purpose:** Children living with disabilities (CLWD) are at high risk of malnutrition but have long been marginalised in malnutrition treatment programmes and research. The 2013 World Health Organization (WHO) guidelines for Severe Acute Malnutrition (SAM) mention disability but do not contain specific details for treatment or support. This study assesses inclusion of CLWD in national and international SAM guidelines.

**Methods:** National and international SAM guidelines were sourced online and via direct enquiries. Eight scoping key informant interviews were conducted with experts involved in guideline development to help understand possible barriers to formalising malnutrition guidance for CLWD.

**Results:** 71 malnutrition guidelines were reviewed (63 national, 8 international). Only 4% (3/71) had a specific section for CLWD, while the remaining lacked guidance on consistently including CLWD in programmes or practice. Only one guideline mentioned strategies to include CLWD during a nutritional emergency. Most (99%,70/71) did not link to other disability-specific guidelines. Of the guidelines that included CLWD, most only discussed disability in general terms despite the fact that different interventions are often needed for children with different conditions. Interviews pointed towards barriers related to medical complexity, resource constraints, epidemiology (*e*.*g*., unrecognised burden), lack of evidence, and difficulty of integration into existing guidelines.

**Conclusion:** Children living with disability are not recognised in most SAM guidelines. Where they are, recommendations are very limited. Better evidence is urgently needed to identify and manage CLWD in malnutrition programmes. More inclusion in the 2022 update of the WHO malnutrition guidelines could support this vulnerable group.

## INTRODUCTION

Globally, some 1 billion people live with a disability, of whom 93 million are children. ^1^ It is also estimated that 47 million children aged under 5 years are suffering from wasting ^2^, and that a significant, but as yet unknown number of children living with disabilities (CLWD) are amongst these millions of children. Disability and malnutrition interact in many ways: feeding problems related to anatomic or motor impairments, nutrient malabsorption or social exclusion are some of the ways in which underlying disability can increase the risk of malnutrition.^3, 4^

A 2018 systematic review found the pooled odds ratio for undernutrition was three times higher for CLWD, compared to non-disabled children (double for stunting and wasting, respectively).^5^ CLWD also have greater risk of adverse outcomes, including death, following treatment for severe acute malnutrition (SAM).^6^

There is an imperative through the UN Convention on the Rights of Persons with Disabilities and the Sustainable Development Goals ^2, 7^ for international actors to commit to greater inclusion of persons living with disabilities in health care and in related sectors such as humanitarian practice initiatives.^8-10^ The 2013 update of the WHO SAM guidelines recognise disability as an underlying factor for malnutrition. However, disability is only briefly mentioned as a possible reason for referral for specialised care and there are no specific recommendations for this vulnerable patient group.^11^ Their needs may differ from other children regarding more complex feeding problems, slower weight gain despite the same treatment, and higher risk of not achieving nutritional recovery. Anthropometry can be more challenging: measuring weight-for-length may be difficult due to spastic contractures (and different target values may apply, *e*.*g*. in children with growth restrictions), and mid-upper arm circumference (MUAC) accuracy may be influenced by differences in muscle mass or body composition ^12^ changes.

There is an evidence gap around the identification and management of malnutrition in CLWD ^3, 4^ and disability can be an exclusion criterion in some studies on malnutrition.^13^ Greater inclusion of disability in nutrition research has been called for numerous times in the academic literature. ^4, 14, 15^ Still, there remain a number of “unexplored opportunities for collaboration” between malnutrition and disability programmes, a need for more political and resource commitment, and a need for malnutrition policies and guidelines to contain detailed guidance specific to CLWD leading to higher visibility of people living with disabilities in front-line practice.^15^

This study aims to provide an overview of the current status of recommendations for CLWD in national and international SAM guidelines, in light of the ongoing 2022 WHO malnutrition guideline update. These new guidelines will influence malnutrition care over the coming decade, and we hope this study will help highlight the need of more inclusivity.

## METHODS

We aimed to identify the most current version of available national and international guidelines focusing on SAM in children aged 0-18 years. These were searched online using manual searching via common search engines, and using resource collections of national health ministries, malnutrition working groups, and international organisations. Contacts of the co-authors, UNICEF regional office teams, and respondents to a call posted on the Emergency Nutrition Network (ENN) global technical forum (https://en-net.org) helped identify additional guidelines. **Table 1** provides details on guideline search strategies and sourcing. The most current final or draft/interim version of national and international protocols were included. Protocols in English, French, Spanish, or Portuguese were reveiwed. Incomplete documents, regional guidelines, or protocols sub-specialising in a certain patient group (e.g. people living with HIV) were excluded. After a full-text review of each guideline, relevant content on disability-specific information, clinical recommendations, monitoring and evaluation indicators, malnutrition prevention strategies and links to other pertinent guidelines were extracted into an electronic database.

**Table 1.**
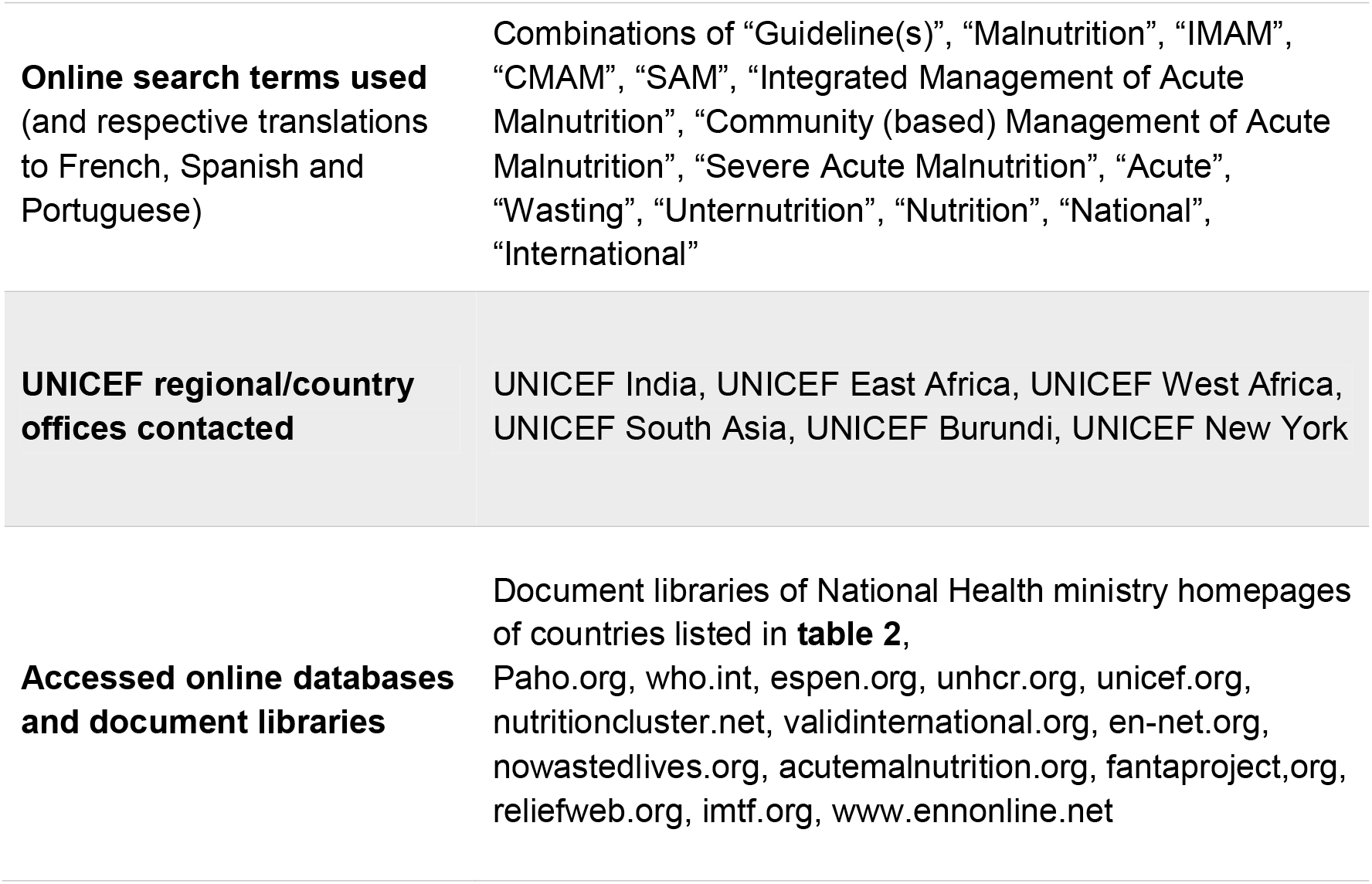
Search terms, contacts and online resources used

Scoping key informant interviews were conducted to understand perceived barriers to formalising guidance specific to malnourished CLWD. Based on previous work, we aimed for 10 participants to reach content saturation.^16^

Interview participants sought were experts on malnutrition involved in malnutrition guideline development in various roles and countries. They were contacted via email using established professional networks, via an ENN forum call, and by snowball sampling.

Informants received an information sheet, signed a consent form, and were sent a previously piloted interview guide. They commented purely in a personal capacity, not representing any institutions or organisations.

Interviews were conducted by the lead author via an online video call and lasted approximately 45 minutes. The interview was recorded, transcribed and anonymised, and participants were offered the transcripts to verify accuracy. Thematic analyses of the transcripts were performed following the steps outlined by Richie and Spencer.^17^

## RESULTS

71 malnutrition guidelines were identified, of which 89% (63/71) were national guidelines from 56 countries (see **Table 2**). For seven countries, more than one guideline was included as separate protocols existed *e*.*g*. for inpatient/outpatient treatment. National guidelines were distributed across the following UNICEF-designated regions: 32% (20/63) from West and Central Africa, 29% (18/63) from Eastern and Southern Africa, 14% (9/63) from South Asia, 14% (9/63) from East Asia and the Pacific, 6% (4/63) from the Middle East and North Africa, and 5% (3/63) from Latin America and the Caribbean. The final 11% (8/71) were international guidelines from one or more NGOs.^18-25^ 75% (53/71) were published in 2013 and later.

**Table 2.**
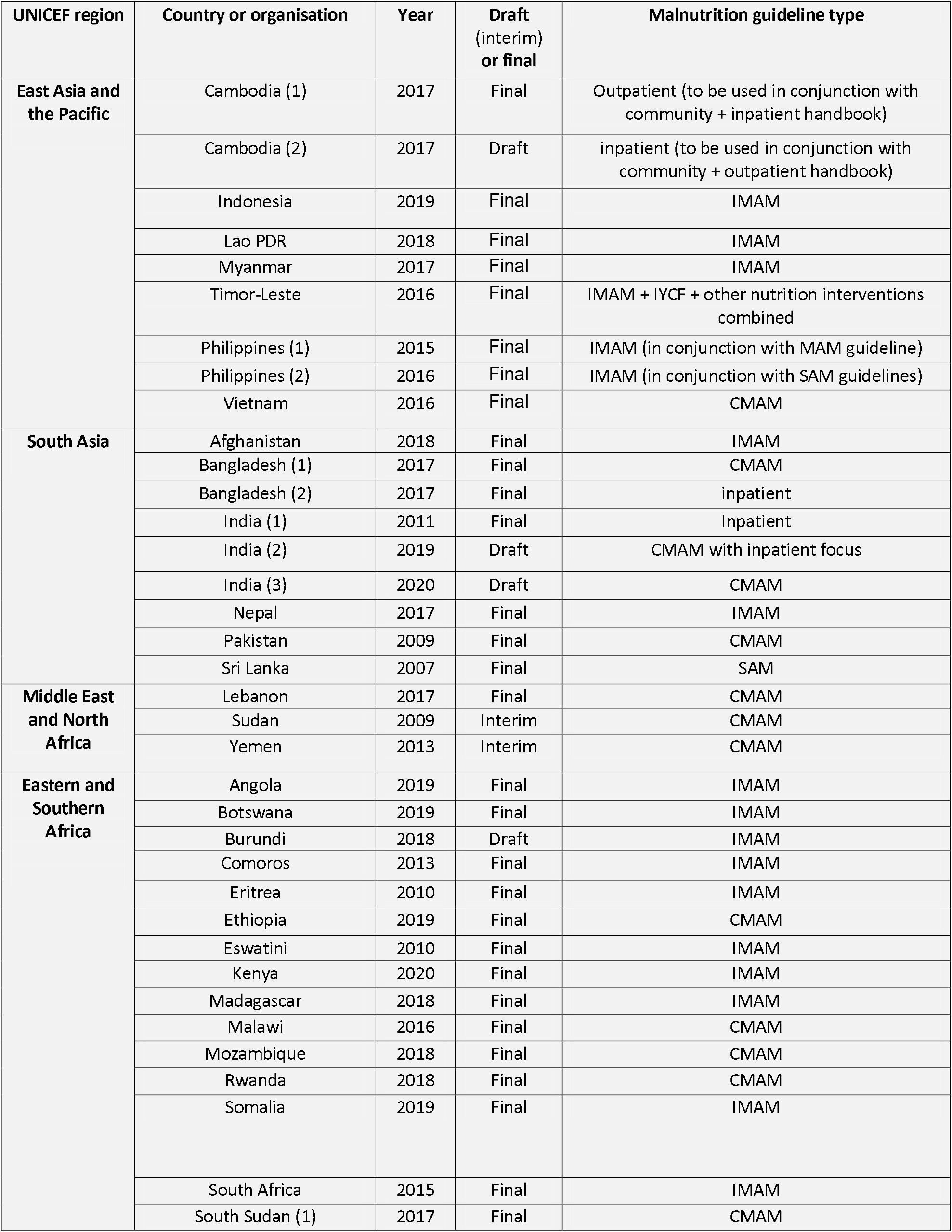
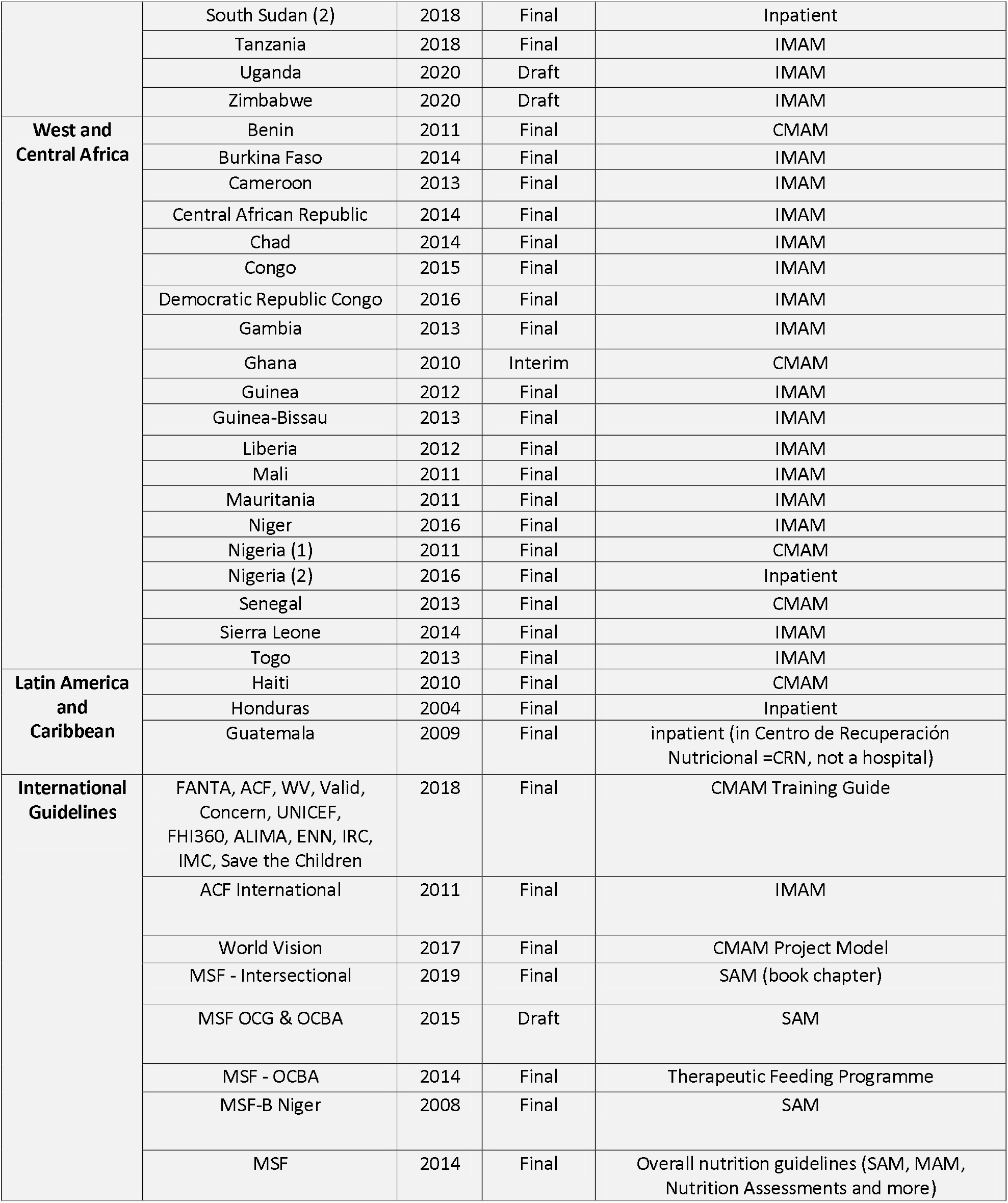
Guidelines included in this review

While 85% of the guidelines (60/71) mention disability, most did so briefly. Only 4% (3/71) have a dedicated section on children living with disabilities. No international guideline is amongst these.

Regular integration of CLWD into the body of the guidelines was also rare. With few exceptions, the extent of referencing disability in guidelines is limited to side notes:

○ 43/60 (72%) note disability as a possible indication for nasogastric tube feeding (e.g. cleft lip/palate)
○ 39/60 (65%) acknowledge disability as a possible reason for treatment failure or non-response
○ 18/60 (30%) suggest disability as a reason for inpatient treatment
○ 6/60 (10%) recognize disability as a risk factor for malnutrition

Cerebral palsy and cleft lip/palate were commonly mentioned disabilities, while some refer broadly to “congenital abnormalities”. Almost no guidelines responded to specific needs of CLWD:

○ 1/60 outlined specific breastfeeding advice for children with cleft lip/palate (Timor-Leste 2016^26^)
○ 1/60 specified a multidisciplinary approach for managing children with cerebral palsy and gave feeding counselling advice (Lebanon CMAM 2017^27^)
○ 18/60 (30%) recommend the “medical referral” of CLWD to a specialist

While 70% (42/60) of guidelines mention the need to identify disability in the physical assessment of a child, this primarily meant listing a box to check “disability yes/no” on a physical assessment form. Only one guideline (Ethiopia CMAM 2019^28^) advises users to “proactively screen for disability”. One other protocol (Timor-Leste 2016^26^) specifies to “also look for developmental delay”.

One protocol (Lebanon 2017^27^) linked the use of their protocol to other disability-specific guidance available (a cerebral palsy feeding and nutrition review^29^). 6/60 protocols (10%) mentioned social or mental health support for CLWD and their families:

○ 1/60 to establish a link with supporting families in nutritional emergency situations
○ 1/60 to prioritise CLWD for psychosocial support
○ 1/60 discussed informing caretakers on realistic outcome expectations (e.g. slower weight gain)
○ 1/60 recommended home visits for CLWD
○ 2/60 encouraged to provide counselling on the nutritional needs of CLWD
○ 3/60 to “refer to appropriate support services” (no specifics)

Very few guidelines (3/60, 5%) include any Monitoring and Evaluation (M&E) indicators specific to disability.

There were four positive examples of more inclusive guidelines that are highlighted in **Table 3**. Three of these contain separate chapters on CLWD and the fourth mentions strategies to support CLWD in the context of a humanitarian emergency.

**Table 3.** Positive examples of guidelines with recommendations for children living with disabilities

|  |  |
| --- | --- |
| **Ethiopia 2019 CMAM<sup>28</sup>** | ○ Specific chapter (“Vulnerable groups”)<br>○ Recognises link between disability and malnutrition<br>○ Proactively screens for disability<br>○ “Offer disability-specific feeding advice” (but does not specify)<br>○ Counselling on realistic outcome expectations<br>○ Regular home visits<br>○ Referral to support services |
| **Lebanon 2017 CMAM<sup>27</sup>** | ○ Specific chapter on CLWD<br>○ Extensive information on cerebral palsy: team treatment approaches (including gastroenterologists, speech therapists, dieticians, nurses, psychologists), feeding advice, providing daily micronutrient supplements<br>○ Link with support structures<br>○ Refers to guidance document on cerebral palsy |
| **Malawi 2016 CMAM<sup>37</sup>** | ○ Specific sub-heading (SAM in CLWD)<br>○ Recognises link between disability and malnutrition<br>○ Proactively look for CLWD<br>○ Proposes substituting RUTF with F100 (e.g. if cleft palate, cerebral palsy)<br>○ Counsel and advise parents about disability<br>○ “Offer disability-specific feeding and treatment”<br>○ “Provide realistic outcome expectations”<br>○ Refer to appropriate services |
| **Timor-Leste 2016 Specific Nutrition Intervention Package Guidelines<sup>26</sup>** | ○ Lists CLWD as vulnerable group in emergencies (but notes that prevalence of malnutrition in this group is unknown)<br>○ In emergencies: give general ration, organize food distribution adapted to their needs, link disabled people with supporting families for joint preparation of meals, health/nutrition/hygiene education, gives feeding advice for children with oral malformations |

**Note:** CMAM = Community-based management of acute malnutrition RUTF = Ready to use therapeutic food

### Perceived barriers to formalising recommendations

Eight malnutrition experts (three female, five male) were recruited for the scoping interviews (zero participation refusals). Participants worked as clinicians, academics, policy makers and/or independent consultants; all had been involved in the development of malnutrition guidelines. Regarding perceived barriers to including CLWD in malnutrition guidelines, five main themes emerged: *Medical expertise, Resources, Epidemiology, Evidence*, and *Guideline Structure*. Within *medical expertise*, subthemes identified were *available diagnostics* and *staff knowledge*.

Regarding medical expertise, the perceived medical complexity, wide range of disabilities and difficulty in developing a guideline that fits all were brought up by several experts (5/8) as a key challenge. As one participant said, *“That’s where the problem starts to get really complicated (…). Is it mental or is it physical? (…) what is it that’s causing the disability? What are the functional outcomes (…)?”*.

Several informants (5/8) mentioned a weak evidence base for recommendations, including around the correct use of anthropometrics (*e*.*g*., special growth charts for CLWD). One participant said, *“I have looked hard enough for it. But (…) I found very little…”*. Another mentioned the difficulties of designing studies to include CLWD as the various disabilities make for a complex and heterogenous study population, with arising data difficult to interpret.

Three participants brought up possible resource constraints, one clinician pointing out that *“They have a much harder time recovering (…), so then they get treated again (…)And they’re in the program for months (…) So, it become very expensive to the program (…)”*.

Epidemiology was discussed by four informants, *e*.*g*. noting that CLWD may not be officially registered (and thus the prevalence remaining unrecognised).

Two participants commented on guideline structures, one saying that recommendations on CLWD may be hard to fit into a space-constrained document. Another discussed the benefits and drawbacks of the creation of many separate guidelines (*e*.*g*., for malnutrition in people living with HIV, in the elderly, in CLWD,…).

## DISCUSSION

This study shows that children living with disabilities are still largely invisible in most national and international malnutrition guidelines. Due to the wide range of disabilities, this can be a complex topic, as some children living with different types of disability need tailored approaches.

A first step in making CLWD visible in guidelines and front-line practice is recommending active screening for disability during case finding and admission to malnutrition treatment programmes, be that inpatient or community-based. Recent work suggests that formal screening tools may help with this: many types of disabilities, especially those that are less severe or less obvious are missed via routine clinical examination alone.^6^ Some disabilities can be challenging to diagnose clinically, and malnutrition treatment failure, relapse cases or malnutrition in older children should prompt investigations towards disability. Technical difficulties in measuring the height or length of children with postural impairments (*e*.*g*., muscle contractures in cerebral palsy) also need to be addressed, for example via alternative indicators.^30^ Not including specific indicators for CLWD during M&E processes perpetuates their invisibility, increasing the likelihood that the scale of the problem will remain difficult to recognise.

Reiterating the human rights imperative of providing every child with the chance to survive and thrive, there are a variety of opportunities for increasing the chances of CLWD to receive optimal care, ranging from awareness of risk and the early identification of CLWD to appropriate treatment, more intensive follow-up, and linking with social, economic and mental health support for the child and family. A small but increasing number of resources exist to support feeding and nutrition in children with disabilities, ranging from models for social support to more clinical guidance.^31-35^

The evidence base relating to malnutrition in CLWD is sparse and needs to be strengthened by acknowledging this as a research priority. Whilst it is good that the 2013 WHO SAM guideline update mentions disability, details in the guideines are very limited and so it is arguably unsurprising that eight years on, there is a continuing lack of tailored recommendations in the protocols reviewed in this study. Many national and international malnutrition guidelines are modelled on the WHO guidelines, and several are based on template guidelines. In the case of the protocols reviewed here, several were derived from the 2011 IMAM Generic Protocol developed by Golden and Grellety.^36^ Changes in these widely used templates towards more inclusion of CLWD, or the creation of new guides to mainstream disability into all aspects of CMAM guidelines could have a significant impact on protocols globally. The upcoming 2022 update of the WHO SAM guidelines is a unique opportunity to create more inclusive guidelines. It offers the opportunity to include disability in the search strategy of the systematic reviews commissioned for evidence collection. Guidelines may consider a distinct section for the treatment of SAM in CLWD to highlight their vulnerability and respond to their needs. Another option may be the integration of CLWD all throughout updated guidelines, as this may still further their inclusion if viewed as an integral part of all treatment programmes rather than a separated patient group. However, the evidence base for such guidance remains sparse.

Recommendations need to go beyond mentioning disability as a side note. There is a need to be specific and detailed to allow health workers without extensive experience in disability to provide optimal care. Stating ‘provide feeding as adequate for the respective condition’ is insufficient without specifying what this would mean for children with different types of disability; there needs to be more detailed ‘how-to’, step-by-step guidance. In order to create this kind of guidance, disability needs to be included in the systematic evidence gathering for malnutrition guideline development and should be highlighted as a research priority.

Integrating disability into M&E and nutrition and coverage surveys is another vital step to ensure the actual caseload of CLWD more visible. National guidelines would benefit from including country-specific information on available support services for CLWD, calling upon the expertise of both national and international advocates and experts. If the scope of the guidelines does not allow for this level of detail, a link to other guidance should be established. The option of entirely separate country-specific protocol documents for malnutrition in CLWD (similar to existing stand-alone guidelines for malnutrition in people living with HIV), a related template guideline, and/or a guide to mainstream disability into national CMAM protocols could be discussed, and may be appropriate for some countries although the merits and drawbacks of these approaches must be considered on a country-by-country basis.

This study has several limitations. First, even if a guideline contains specific recommendations for CLWD, this does not necessarily translate into practical, effective implementation, which is likely to vary in different settings. Second, not all existing national and international guidelines could be identified for inclusion in this study. Nevertheless, the 63 national guidelines included cover the greater part of countries with a high burden of malnutrition, thus, we believe, valid conclusions can still be reached. Finally, interviews were scoping interviews to help with initial undertstanding of the issues. Future work would include a wider range of key informants, *e*.*g*. government officials involved in guideline development and front line staff working in implementation and management of malnourished CLWD.

In conclusion, children living with disabilities are still largely marginalised in malnutrition guidelines intended for all children. Though recognition is an important first step, much work needs to be done to expand guidance on the specifics of identification and management of CLWD. We hope our findings can help raise awareness of the need for a commitment towards greater visibility and inclusion of malnourished children living with disabilities.

## Data Availability

Data are stored in the LSHTM Data Compass repository and are available upon reasonable request.

https://doi.org/10.17037/DATA.00002513

## DATA AVAILABILITY STATEMENT

Data are stored in the LSHTM Data Compass repository and are available upon reasonable request.

## ETHICS STATEMENT

This study was approved by the London School of Tropical Medicine (LSHTM) ethics committee (Reference Number 21844).

## ACKNOWLEDGEMENTS

We sincerely thank everyone helping to source the guidelines, and all the interviewees for taking time to share their insights with us.

## COMPETING INTERESTS

none declared.

## FUNDING

ME (part-funded), CA, NL and MM were funded by Irish Aid and the Eleanor Crook Foundation. MK also acknowledges the Eleanor Crook Foundation for funding support.

## CONTRIBUTORS

MK and ME had the initial idea for this paper. Other authors contributed to a different earlier version of the guideline review project. MK led the design process. ME did the literature and guideline review. Interviews were conducted, transcribed and analysed by ME. The first draft of the paper was written by ME, which was reviewed and revised by all coauthors.

## COREQ Checklist

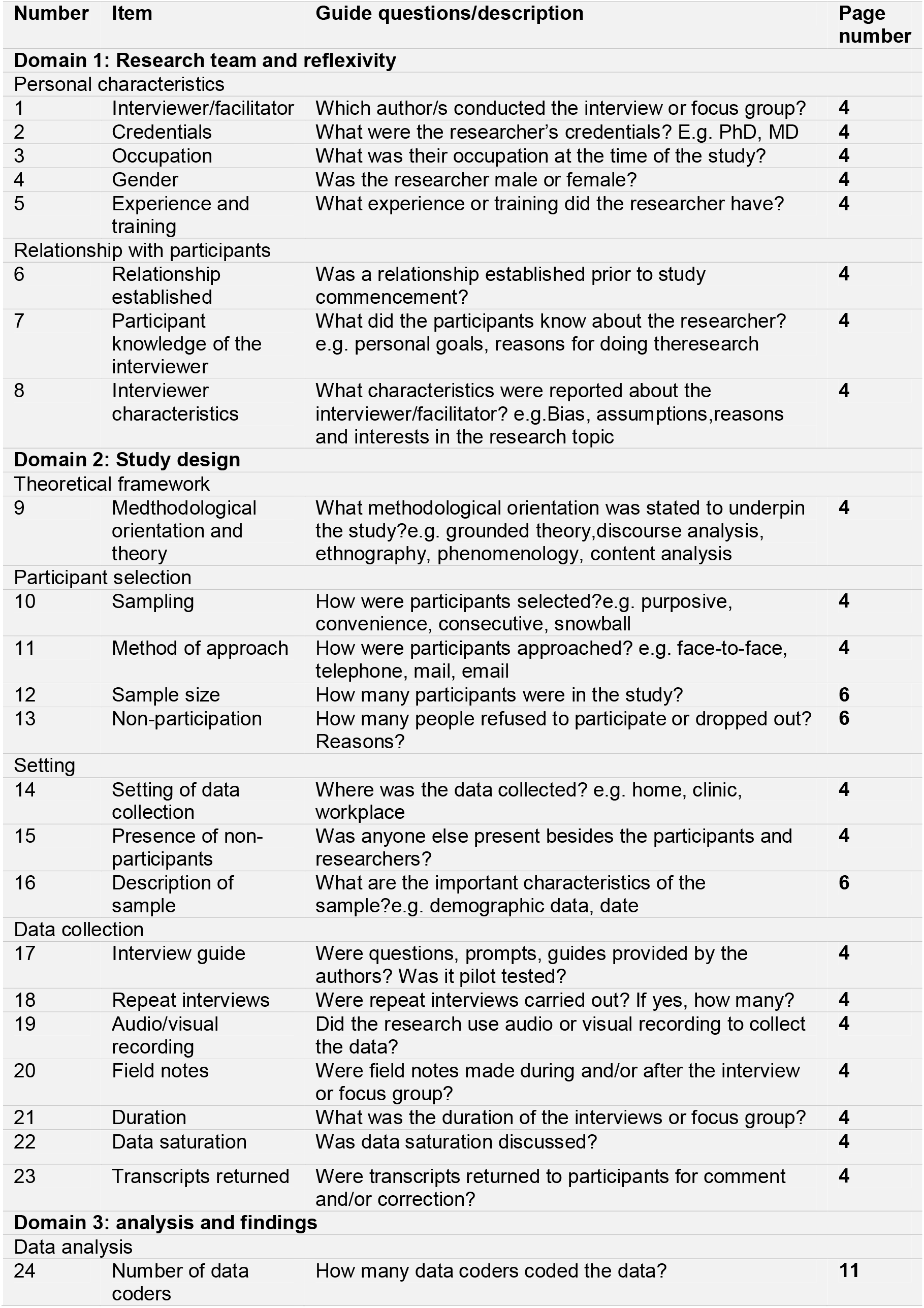
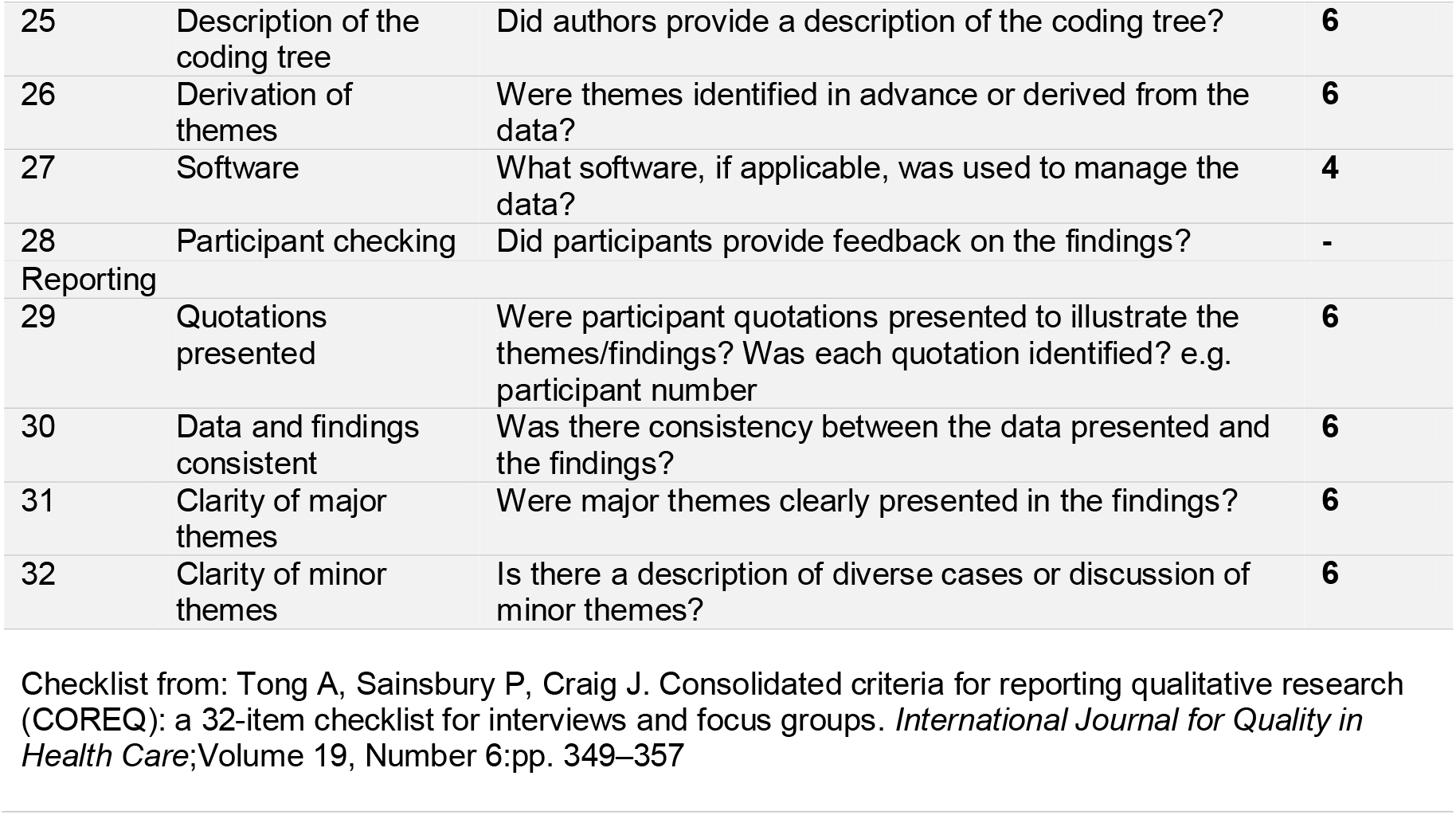

## Notes

### Competing Interest Statement

The authors have declared no competing interest.

### Summary of Updates

The funding statement and the first author's address have been amended. A spelling mistake in one co-author's name (NL) has been corrected.

## REFERENCES

1. The World Bank. World report on disability. Malta: World Health Organization. 2011.

2. United Nations. The Sustainable Development Goals Report 2020 2020 [Available from: https://unstats.un.org/sdgs/report/2020/.

3. Kuper H, Nyapera V, Evans J, et al. Malnutrition and childhood disability in Turkana, Kenya: Results from a case-control study. PloS one. 2015;10(12):e0144926.

4. Groce NE, Kerac M, Farkas A, et al. Inclusive nutrition for children and adults with disabilities. The Lancet Global Health. 2013;1(4):e180–e1.

5. Hume-Nixon M, Kuper H. The association between malnutrition and childhood disability in low-and middle-income countries: systematic review and meta-analysis of observational studies. Tropical Medicine & International Health. 2018;23(11):1158–75.

6. Lelijveld N, Groce N, Patel S, et al. Long-term outcomes for children with disability and severe acute malnutrition in Malawi. BMJ global health. 2020;5(10):e002613.

7. The United Nations. Convention on the Rights of Persons with Disabilities. 2006.

8. Akerkar S, Bhardwaj R. Good practice guide: embedding inclusion of older people and people with disabilities in humanitarian policy and practice. Age and Disability Consortium (ADCAP). 2018:1–160.

9. UNICEF. Core Committments for Children in Humanitarian Action 2020 [Available from: https://www.unicef.org/emergencies/core-commitments-children.

10. Stough LM, Kang D. The Sendai framework for disaster risk reduction and persons with disabilities. International Journal of Disaster Risk Science. 2015;6(2):140–9.

11. World Health Organization. Updates on the management of severe acute malnutrition in infants and children. Geneva: World Health Organization. 2013.

12. McDonald CM, Abresch-Meyer AL, Nelson MD, et al. Body mass index and body composition measures by dual x-ray absorptiometry in patients aged 10 to 21 years with spinal cord injury. J Spinal Cord Med. 2007;30 Suppl 1(Suppl 1):S97–S104.

13. Lelijveld N, Kerac M. A holistic approach to malnutrition follow-up care. Journal of Public Health and Emergency. 2017;1(4).

14. Kerac M, Postels DG, Mallewa M, et al. The interaction of malnutrition and neurologic disability in Africa. Seminars in pediatric neurology. 2014;21(1):42–9.

15. Groce N, Challenger E, Berman-Bieler R, et al. Malnutrition and disability: unexplored opportunities for collaboration. Paediatrics and international child health. 2014;34(4):308–14.

16. Read S, McGrath M. Community management of uncomplicated malnourished infants under six months old: barriers to national policy change. Field Exchange 57. 2018:27.

17. Ritchie J, Spencer L. Qualitative data analysis for applied policy research. The qualitative researcher’s companion. 2002;573(2002):305–29.

18. Médecins sans Frontières. Clinical Guidelines 2020 [Available from: medicalguidelines.msf.org.

19. Médecins sans Frontières. Nutrition guidelines 2014 [Available from: https://bibop.ocg.msf.org/docs/16/L016NUTM01E-P_Nutrition-2014.pdf.

20. Médecins sans Frontières OCG & OCBA. Protocols for Management of Nutritional Support in Children with Severe Acute Malnutrition. Internal Guideline - Pilot Version 2015 [Available from: https://bibop.ocg.msf.org/docs/16/L016NUTM19E-E_PEDNUT-Nut-EN_2015.pdf.

21. Médecins sans Frontières OCB Niger. Protocole Nutritionnel - Médical Nouveau-nés et nourrissons 2008 [Available from: No URL available (internal document).

22. Médecins sans Frontières OCBA. Therapeutic feeding programme - nutritional and medical protocol. All age groups. 2014 - revised 2015 [Available from: No URL available (internal document).

23. ACF International. Guidelines for the integrated management of severe acute malnutrition: in- and out-patient treatment 2011 [Available from: https://www.actionagainsthunger.org/publication/guildines-integrated-management-severe-acute-malnutrition-and-out-patient-treatment.

24. FANTA III UNICEF Concern et al. Training Guide for Community-Based Management of Acute Malnutrition (CMAM) 2018 [Available from: https://www.fantaproject.org/focus-areas/nutrition-emergencies-mam/cmam-training.

25. World Vision. Community-Based Management of Acute Malnutrition (CMAM) - Project Model 2017 [Available from: https://www.wvi.org/nutrition/project-models/cmam.

26. Ministry of Health Timor-Leste. Specific Nutrition Intervention Package (SNIP) Guidelines 2016 [Available from: No URL available, obtained through contacts.

27. Republic of Lebanon Ministry of Public Health. Management of Acute Malnutrition - National Guidelines 2017 [Available from: No URL available, obtained through contacts.

28. Government of Ethiopia Federal Ministry of Health. National Guideline for the Management of Acute Malnutrition in Ethiopia 2019 [Available from: No URL available, obtained through contacts.

29. Ferluga ED, Archer KR, Sathe NA, et al. Interventions for feeding and nutrition in cerebral palsy. Comparative Effectiveness Reviews. 2013;94.

30. Yousafzai AK, Filteau SM, Wirz SL, et al. Comparison of armspan, arm length and tibia length as predictors of actual height of disabled and nondisabled children in Dharavi, Mumbai, India. European Journal of Clinical Nutrition. 2003;57(10):1230–4.

31. Zuurmond M, O’Banion D, Gladstone M, et al. Evaluating the impact of a community-based parent training programme for children with cerebral palsy in Ghana. PloS one. 2018;13(9):e0202096.

32. Holt International. Holt International’s Feeding and Positioning Manual: Guidelines for Working with Babies and Children 2019 [Available from: https://www.holtinternational.org/about/child-nutrition/feeding-and-positioning-manual/.

33. LSHTM ICD CBM CSF Hambisela. Getting to know cerebral palsy: Working with parent groups – a training resource for facilitators, parents, caregivers, and persons with cerebral palsy [Available from: https://www.lshtm.ac.uk/sites/default/files/2019-06/Getting-to-know-cerebral-palsy-english.pdf.

34. MCAI (Maternal & Childhealth Advocacy International). International Maternal & Child Health Care: A practical manual for hospitals worldwide: Radcliffe Publishing Ltd.; 2014.

35. SPOON. Count Me In http://www.spoonfoundation.org/what-we-do/our-approach/count-me-in/; 2019.

36. Golden M, Grellety Y. Golden MH Grellety Y, Integrated Management of Acute Malnutrition (IMAM) Generic Protocol ENGLISH version 6.6.2 2011 [Available from: https://www.researchgate.net/publication/292131715_Golden_MH_Grellety_Y_Integrated_Management_of_Acute_Malnutrition_IMAM_Generic_Protocol_ENGLISH_version_662.

37. Government of Malawi Ministry of Health. Guidelines for community-based management of acute malnutrition 2016 [Available from: https://www.fantaproject.org/sites/default/files/resources/Malawi-CMAM-Guidelines-Dec2016.pdf.

